# Human Population Density Drives Pneumococcal Disease Risk Via Density-Dependent Serotype Transmission

**DOI:** 10.64898/2025.12.05.25341703

**Authors:** Benjamin J. Metcalf, Kristofer Wollein Waldetoft, Bernard W. Beall, Sam P. Brown

## Abstract

In human infectious diseases, the likelihood of infection in a susceptible individual is generally assumed to be dependent on the frequency of infected individuals in the population (higher infection prevalence means more risk of infection per contact) and not dependent on population density. In this investigation we use CDC Active Bacterial Core surveillance data on over 34 million US citizens to test for density-dependent transmission in the pneumococcus. We show that 1) *Streptococcus pneumoniae* (Spn) transmission is dependent on host population density and 2) serotypes with shorter carriage durations and higher invasiveness are more likely to transmit in more populated areas. These results have important implications for pneumococcal epidemiology, given substantial variation in human population density, and the connection of carriage duration with critical infection properties including antimicrobial resistance, recombination, invasiveness, and serotype prevalence.

## Introduction

Infectious diseases are generally dependent on social interactions among their hosts. Contact-mediated infection requires susceptible individuals to encounter infected individuals (Borremans et al., 2017). Intuitively, the force of infection *λ*, the rate of infection experienced by susceptible individuals, will depend on 1) the density of hosts (more individuals mean more social contacts), and 2) the frequency of infected individuals in the population (higher infection prevalence means more risk of infection per contact). In mathematical models of infectious disease dynamics, these two dependencies have become associated with two limit-case models (Begon et al., 2002), termed ‘density-dependent transmission’ and ‘frequency-dependent transmission’ (Figure 1).

**Figure 1:**
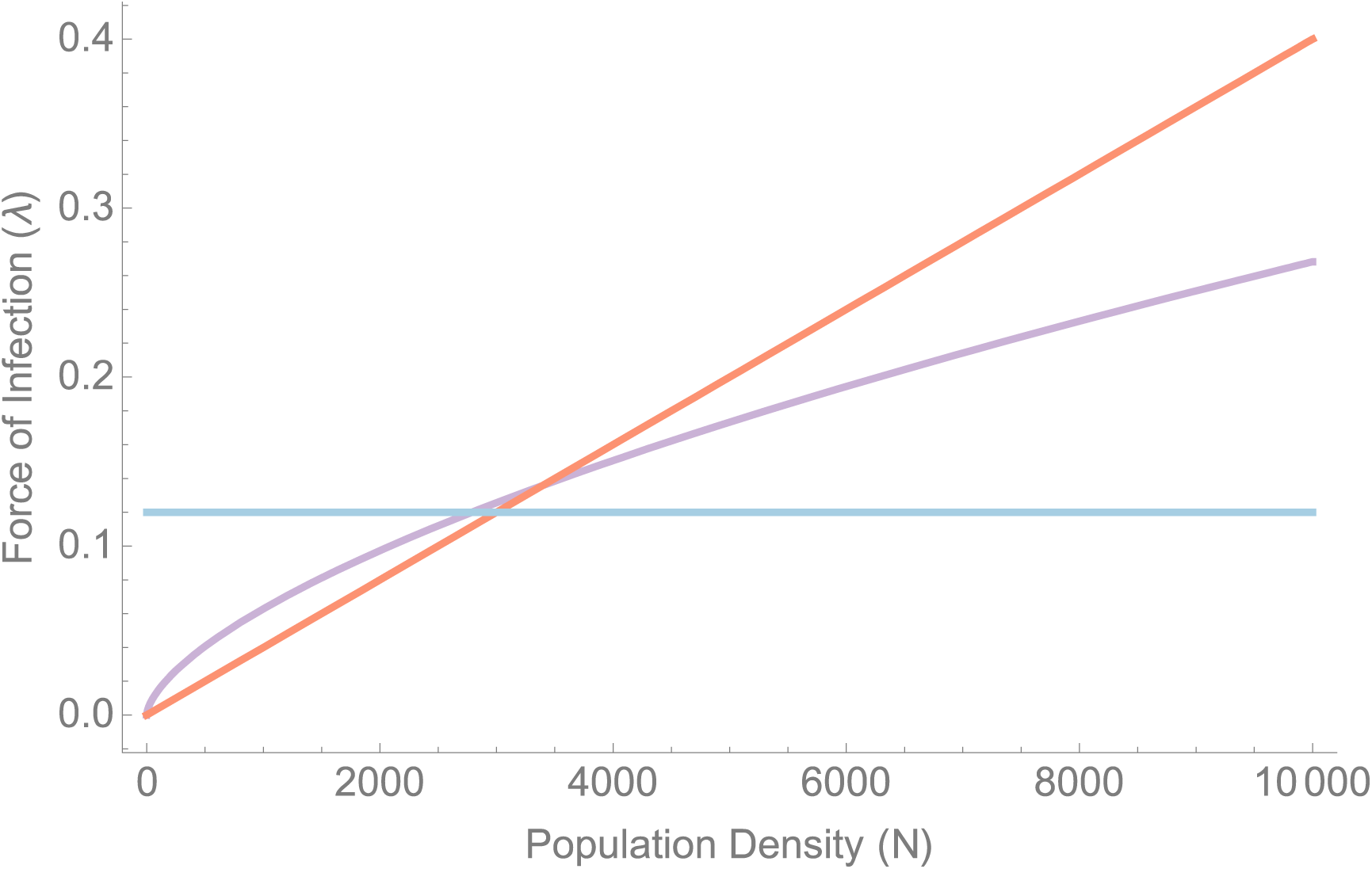
**Models for the force of infection *λ* as a function of population density *N.*** The blue line defines a frequency-dependent contact structure (*λ* = *κI*, where *I* is the frequency of infecteds and *κ* is a rate constant) that is independent of total population density *N*. The red line defines a density-dependent contact structure (*λ* = *κNI*, where *NI* defines the total density of infecteds in the population). The purple line defines a power contact structure (λ = *κN^r^I,* contact rate = *κN^r^*) and represents a ‘hybrid’ of frequency- and density-dependent limit cases (when *r =* 0 we recover the pure frequency-dependent model, and when *r =* 1 we recover the pure density dependent model).

Under the frequency-dependent limit case, *λ* is proportional to the *frequency* of infecteds in the population, *I* (blue line, Fig. 1). For human communicable diseases, it has been generally assumed that interaction patterns don’t scale strongly with density and that a frequency-dependent contact structure is sufficient (Grassly and Fraser, 2008, Ferrari et al., 2011) since whether you live in a small community or a mega-city, class-room sizes and workplaces where transmission takes place tend to be on a similar scale. In agreement with this interpretation, an analysis of UK measles time-series data in the pre-vaccination era supported a frequency-dependent model (Bjornstad, 2002).

Under the density-dependent limit case by contrast, *λ* is proportional to the *density* of infected individuals, *IN* (red line, Figure 1), where *N* is the total population density. Wildlife ecologists have demonstrated the importance of density-dependent transmission effects especially in the context of pathogen persistence in wildlife reservoirs and control of invasive species (Cross et al., 2013, Cross et al., 2010, Berthier et al., 2000). Accounting for host population density has also been an area of interest for human viral respiratory pathogens like influenza and, more recently, SARS-CoV-2 though the results have been mixed. Investigations into the relationship between influenza and population density have yielded inconclusive results (Garrett, 2007, Mills et al., 2004, Nishiura, 2008, Chandra, 2013) while recent investigations into the spread of SARS-CoV-2 have identified an association between transmission and population density (Sy et al., 2021).

The two models described above can be generalized as limit cases of a broader model, for example by introducing a scaling parameter *r* (purple line, Figure 1), allowing the impact of density to plateau at higher densities (Borremans et al., 2017). In support of this model, work by Cross *et al*. found that *Brucella abortus* transmission in elk was dependent on density but that the contact network of elk herds followed a hybrid model that saturated at higher population densities (Cross et al., 2013, Cross et al., 2010).

With respect to our focal bacterial pathogen *Streptococcus pneumoniae* (Spn), like other respiratory pathogens, we find that prior epidemiological models overwhelmingly rely on a purely frequency-dependent contact function (Fig 1, blue line). In a structured literature review (Supplement), we identified only a single article that assumed density-dependent transmission and that investigation was focused on pneumococcal spread within households (Melegaro et al., 2004).

While Spn can cause illness (O’Brien et al., 2009), it usually colonizes the nasopharynx asymptomatically (referred to as carriage). Symptomatic Spn usually presents as non-invasive diseases like otitis media and non-bacteremic pneumonia, but, on rare occasions, it can also spread into normally sterile sites and cause severe infections known as invasive pneumococcal disease (IPD). Epidemiological studies of pneumococcal disease are typically organized by serotype, with each serotype distinguished by an immunologically distinct polysaccharide (PS) capsule. Sometimes, especially in older studies, serogroup (a more general category of related but distinct serotypes) is reported instead of serotype. PS capsule is the primary virulence factor of Spn and influences several traits including the duration of carriage, invasiveness (propensity to cause invasive disease), and carriage prevalence in a population (Sleeman et al., 2006, Chaguza et al., 2020). Over the first half of the 20th century, the majority of Spn adult invasive disease was attributable to serogroups 1, 2, 3, and 5 despite them being rarely identified in carriage (Feikin and Klugman, 2002). This subset is sometimes referred to as the ‘epidemic’ serogroups due to their propensity to cause outbreaks in crowded environments (Feikin and Klugman, 2002). This association with crowding suggests that some serotypes with low overall carriage rates may thrive in highly dense and crowded populations. While serogroups 1, 2 and 5 are now largely eradicated thanks to vaccine and antibiotic interventions, there remains substantial variation in carriage duration among currently circulating serotypes (Sleeman et al., 2006).

In this investigation, we use extensive CDC Active Bacterial Core surveillance (ABCs) data to test for density-dependent transmission in the pneumococcus. We hypothesize that short carriage duration serotypes can only persist in dense human populations, as they rely on a high frequency of transmission opportunities in their short duration of carriage. We test this hypothesis using IPD data acquired through the ABCs program which conducts active laboratory and population-based surveillance for invasive Spn infections in a population of more than 34.4 million persons. We show that 1) pneumococcal transmission is both density- and frequency-dependent, and 2) virulent, short carriage duration serotypes are enriched in the densest human populations.

## Results

### Population Density is Correlated with Serotype Carriage Duration and Invasiveness

To approach our hypothesis that shorter serotype carriage duration is associated with higher human population density, we begin by characterizing the average human population density associated with each Spn serotype, by combining CDC and census data. The average host population density per serotype is calculated from either census tract or zip code data associated with each individual IPD case report (see Methods for more details). In Fig. 2 we see significant 12-fold variation in serotype-specific human density (ANOVA F(55, 9978) = 9.375, p-value = 2e-16), ranging from dense-population 7F (average population density of 15,037 people per square mile (PPSM), greater than the density of Washington DC) through to low density 23F (average population density of 1,224 PPSM, less dense than an average suburban neighborhood (Airgood-Obrycki and Rieger, 2019)). Across all census tracts with an IPD case, the average population density is 4,502 PPSM, which is not significantly different from the mean population density of 4,600 PPSM across the total surveillance population (Kolmogorov-Smirnov test, p-value = 0.974).

**Figure 2:**
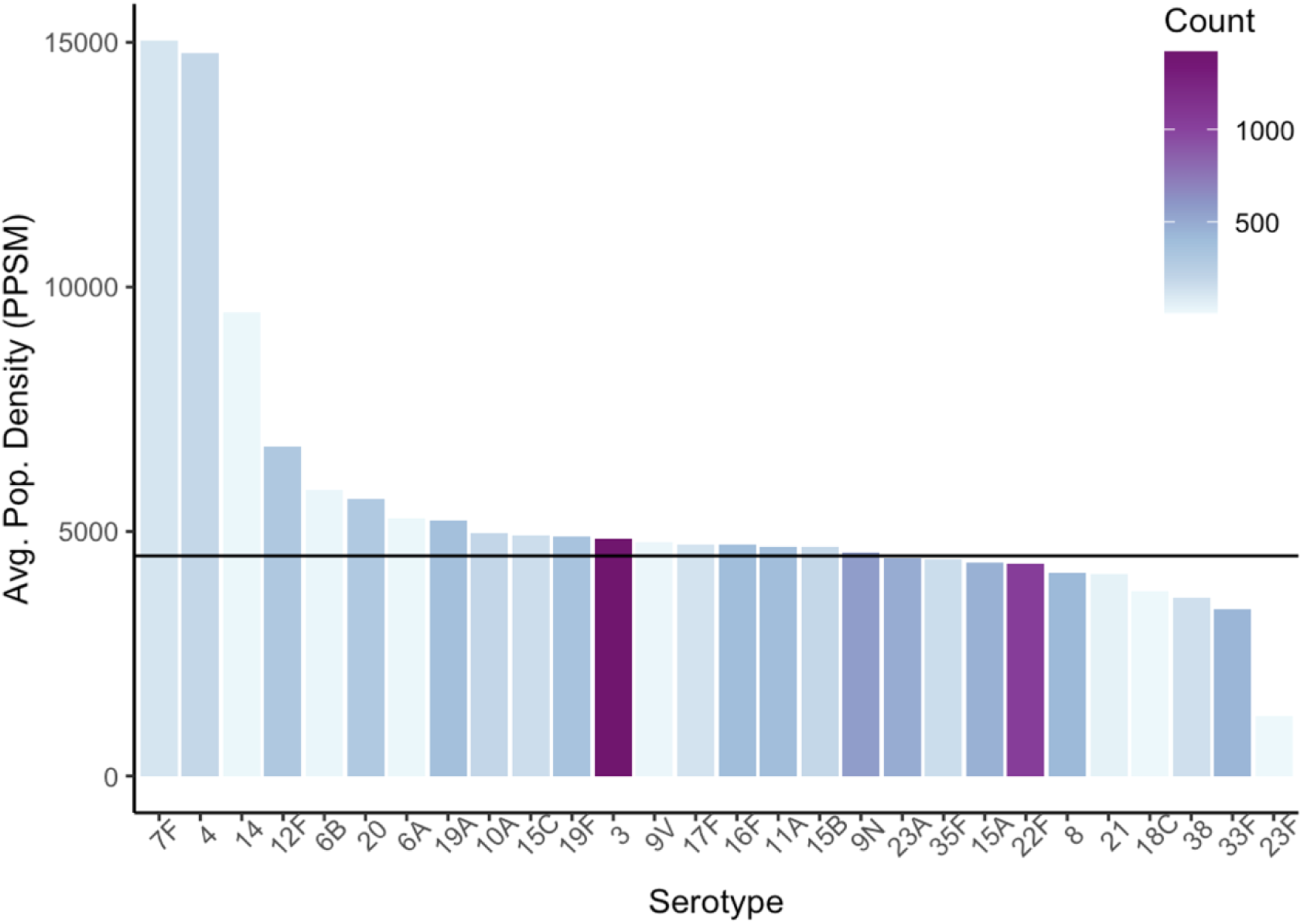
Average serotype population density varies widely across serotypes. Bar chart of average host population density by serotype where population density is measured in persons per square mile (PPSM). The bar color encodes the number of IPD cases for each serotype with the darker color representing higher case counts. Horizontal black line: average population density across all census tracts with IPD cases regardless of serotype.

Given the variation in average population density we see across serotypes, we now ask whether there are bacterial attributes associated with this variation. We focus on two life-history traits, carriage duration and invasiveness, as they have been shown to be serotype-dependent (Sleeman et al., 2006). We first look at the association between the average population density of serotypes acquired from invasive cases with their respective carriage durations (Fig. 3A). A significant inverse correlation between average population density and carriage duration is evident whether we use census tracts (Kendall Tau = -0.348, p-value = 0.00961; Figure 3A) or ZIP codes (Kendall Tau = -0.385, p-value = 0.00415; Fig. S1A) as geographical units of measure. We also examined the relationship between population density and serotype invasive attack rate (Fig. 3B). Here we found a positive relationship that was significant for both census tracts (Kendall Tau = 0.328, p-value = 0.011; Figure 3B) and ZIP codes (Kendall Tau = 0.256, p-value = 0.0476; Figure S1B) as geographical units of measure. These relationships indicate that shorter duration and more highly invasive serotypes are more likely to cause IPD in more dense environments relative to their longer duration and less invasive counterparts.

**Figure 3:**
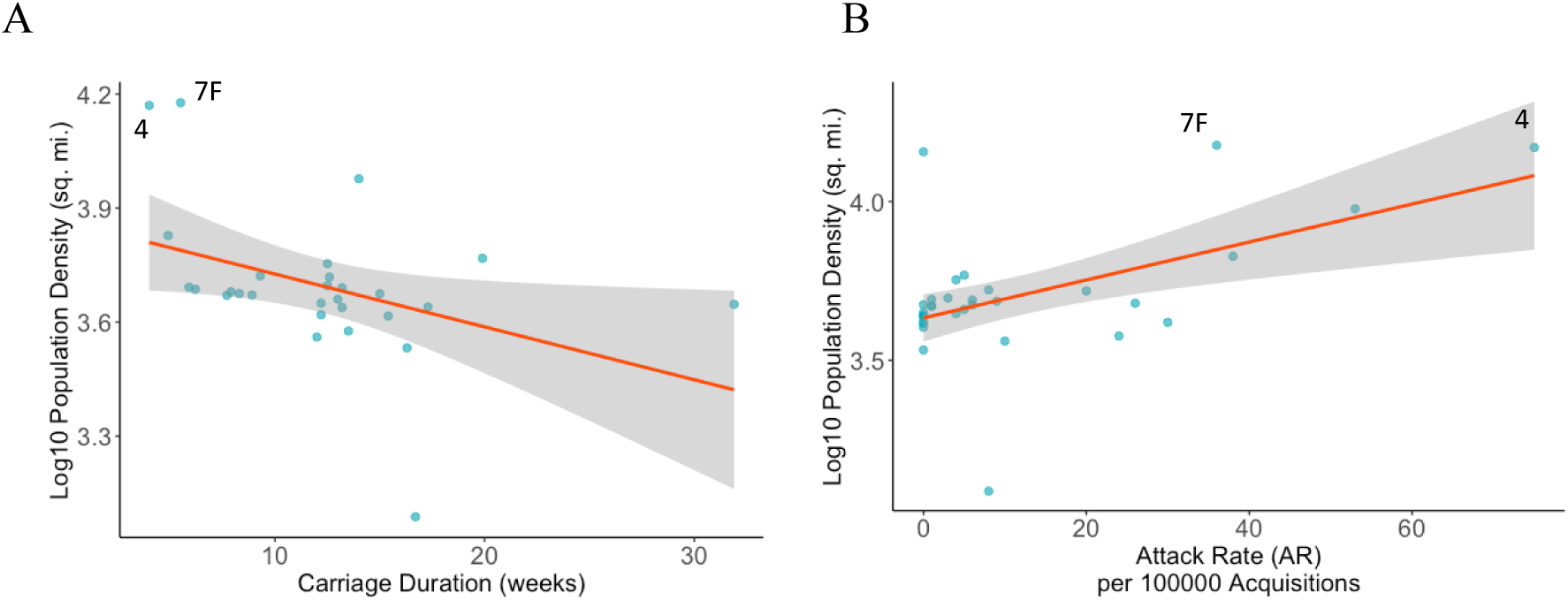
Average serotype population density has a negative correlation with carriage duration and a positive correlation with invasive attack rate (AR). (A) Serotype carriage duration (blue dots) against log10 average serotype population density (measured using census tracts) yields a significant inverse association (Kendall Tau = -0.348, p-value = 0.00961). (B) Invasive AR (blue dots) against log10 average serotype population density (measured using census tracts) yields a significant positive association (Kendall Tau = 0.328, p-value = 0.011). Two examples of short carriage duration, high invasive serotypes (4 and 7F) are labeled in each plot.

### Pneumococcal transmission is both frequency- and density-dependent

The results in Fig 3 support our hypothesis that shorter carriage duration strains (e.g. 4 and 7F) are selected for in more dense environments, and more generally indicate the presence of at least partially density-dependent transmission in the pneumococcus. In order to quantify the extent of density-dependence, we now turn to an explicit model-based approach.

We begin with the compartmental epidemiological model developed previously (Metcalf et al., 2023), tracking the prevalence of susceptible, carriage, invasive disease and recovered cases (*S, C, I, R,* respectively, see methods). On this parameterized baseline, we modify the transmission term to allow for intermediate degrees of density versus frequency dependence. Specifically, we follow two distinct forms of interpolation between the frequency- and density-dependent limit cases described in Figure 1. Note that both interpolation models allow either limit case (pure frequency- or density-dependence) to be supported by data. First, we assess the established power-law interpolation (transmission coefficient *β*(*b,r*) = *bN^r^*, purple line in Figure 1, (Borremans et al., 2017). Second, we assess a piecewise function 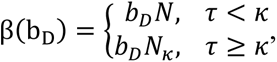 where 𝜏 is serotype carriage duration and 𝜅 is a threshold carriage duration that marks the transition point from density- to frequency-dependent transmission. The piecewise cutoff threshold 𝜅 would naturally be defined in terms of population density instead of carriage duration but is required to be in terms of carriage duration as a constraint of our model fitting procedure (see methods). In the methods we derive a relationship between carriage duration 𝜏 and population density *N* (methods equation 8) that allows us to represent the cutoff threshold 𝜅 in terms of population density (parameter 𝑁_𝜅_). At population density 𝑁_𝜅_, the transmission coefficient *β* transitions to frequency-dependence where *β* is fixed at the threshold value (𝑏_𝐷_𝑁_𝜅_) and invariant across higher population densities.

Along with population density, the IPD case data acquired from ABC surveillance also allows us to calculate invasive incidence for each serotype. While our compartmental model was not specifically designed to model invasive incidence, we can use the incidence data as an additional constraint when estimating the population density model parameters. In the methods we derive a mathematical relationship between carriage duration 𝜏 and invasive incidence (methods equations 7). Functions for both population density and invasive incidence are then used for model fitting and parameter estimation (see methods).

In addition to fitting the relevant force of infection parameters (*b* and *r* for the power law function, and 𝑏_𝐷_, and 𝜅 for the piecewise function), we also fit the duration of immune protection *f*. All other model parameters are obtained from previous work (see methods). Parameter fits under the two force of infection models are presented in Table 1, and the corresponding transmission coefficient *β* as a function of density is presented in Fig. 4.

**Figure 4:**
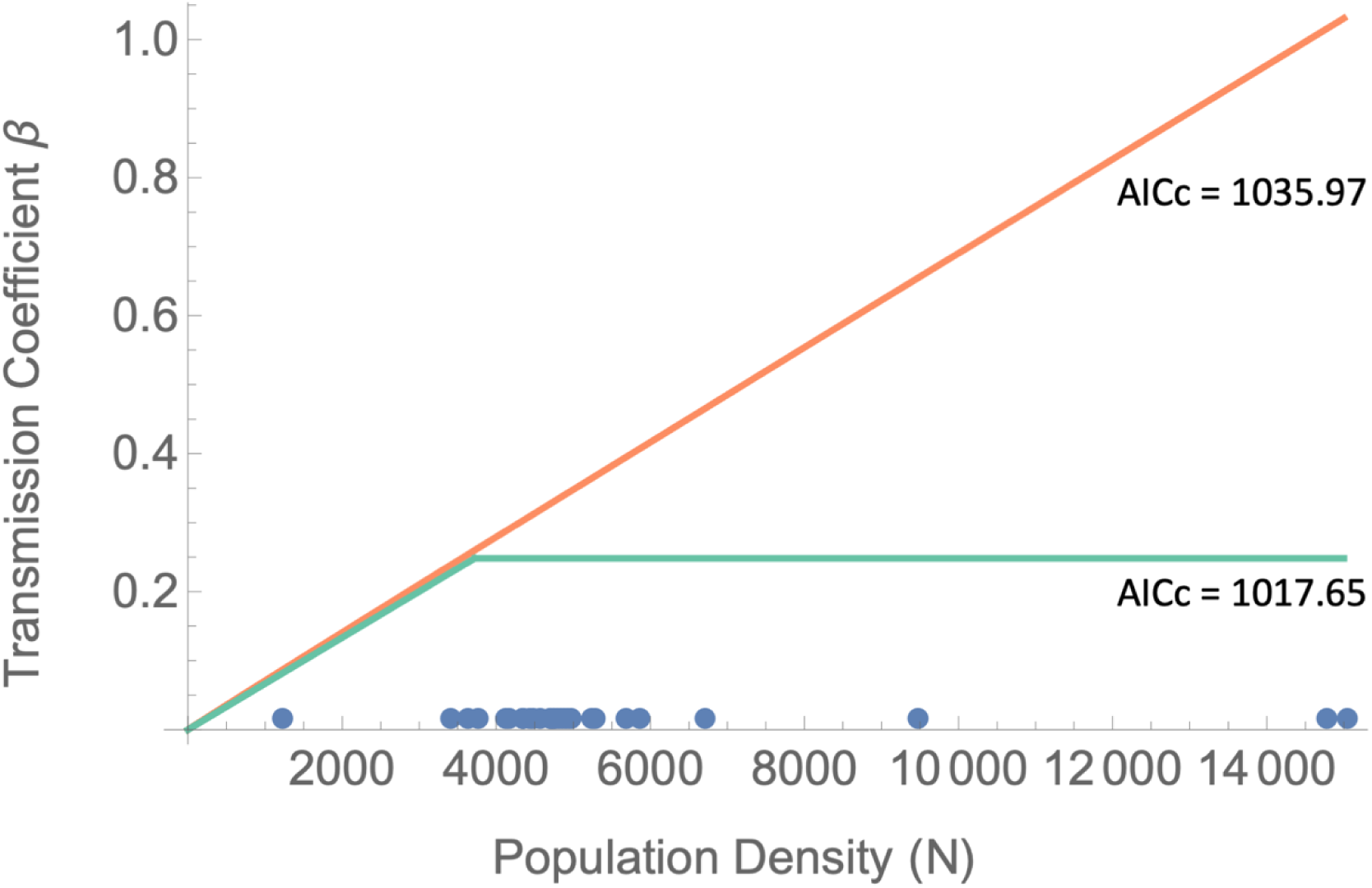
Both the fitted power and piecewise contact rate models indicate that Spn transmission is density dependent. The fitted power contact rate model transmission coefficient *β* (orange line, *β*(*b,r*) = *bN^r^)* where *r* = 0.988 and *b* = 7.69x10-5 against population density *N*. The fitted piecewise contact rate model transmission coefficient *β* (green line, 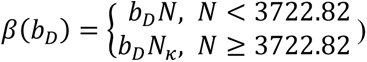 where *b_D_* = 6.67x10-5 against population density *N*. The 𝑏_𝐷_𝑁_𝜅_, 𝑁 ≥ 3722.82 population density threshold of 3722.82 is calculated by plugging the fitted *κ* = 11.07 value into the function for population density (methods equation 8) and using the average per capita invasive incidence of 7.69e-7 cases/week. Both models have a non-constant relationship with population density suggesting that Spn spreads through density-dependent transmission at least in lower population density regions. Model comparisons are made using AICc scores (Burnham et al., 2011). For comparison, the pure density- and frequency-dependent limit cases had AICc scores of 1032.78 and 1222.08 respectively. Blue dots along x-axis represent average serotype population density.

**Table 1:** Fitted coefficient values for parameters used in the power law and piecewise contact rate models.

| Power Law Contact Rate Model |  |  | Piecewise Contact Rate Model |  |  |
| --- | --- | --- | --- | --- | --- |
| Parameter | Definition | Fitted Coefficient Value (95%CI) | Parameter | Definition | Fitted Coefficient Value (95%CI) |
| $r$ | Contact rate exponent | 0.988<br>(0.709 - 1.27) | $\kappa$ | Carriage duration threshold | 11.07<br>(8.52 - 13.62) |
| $b$ | Transmission rate | $7.69 \times 10^{-5}$<br>$(-8.99 \times 10^{-5} - 2.44 \times 10^{-4})$ | $b_D$ | Density-dependent transmission rate | $6.67 \times 10^{-5}$<br>$(4.11 \times 10^{-5} - 9.23 \times 10^{-5})$ |
| $f$ | Duration of immune protection | 236.91<br>(190 - 284) | $f_P$ | Duration of immune protection | 229<br>(189 - 269) |

We estimate the piecewise threshold constant *κ* to be 11.07 (95%CI: 8.52 to 13.62) meaning that Spn transmission is density-dependent for carriage durations less than 11.07 weeks and frequency-dependent otherwise. Plugging this value into our function for population density (methods equation 8) and using the average per capita invasive incidence of 7.69e-7 cases/week, we find the population density threshold 𝑁_𝜅_ is 3722.82 (PPSM). Thus, the frequency-dependent transmission coefficient *β*, calculated by multiplying 𝑁_𝜅_ by *b_D_*, is 0.248 in line with previous work (Domenech de Cellès et al., 2011, Melegaro et al., 2004).

Table 1 and Figure 4 show that both models support density dependence in Spn. Among the two model structures, the piecewise model has greater data support (delta AICc = 18.32, implying a relative likelihood of 9509.06 (Burnham et al., 2011)). For comparison, setting the power law exponent *r* to 0 and fitting a purely frequency-dependent transmission model, we find substantially weaker support (delta AICc = 1222.08, relative likelihood of 2.46e44), allowing us to reject the standard epidemiological assumption of no density dependence.

The piecewise model suggests that Spn transmission is dependent on density below a critical density threshold of 3722.82 PPSM, whereas above this threshold a purely frequency-dependent behavior is supported. Plots of the fitted population density and invasive incidence equations for the piecewise contact rate model are provided in Figures 5 and S2 respectively. The plot of invasive incidence curves shown in Figure S2 also provides an opportunity to check for the internal consistency of the piecewise model fit. If our model design is correct, then when we set population density *N* to be the threshold value 𝑁_𝜅_ for the invasive incidence function (equation 7), the frequency- and density-dependent transmission terms *β* will be identical. Thus, the two individual functions should connect at the piecewise threshold point and create a continuous curve. The plots shown in Figure S2 represent the relationship between carriage duration and invasive incidence across the full range of population densities observed in our data (1224.26 PPSM to 15037.27 PPSM). Plots of the power law model are shown in Figure S3.

**Figure 5:**
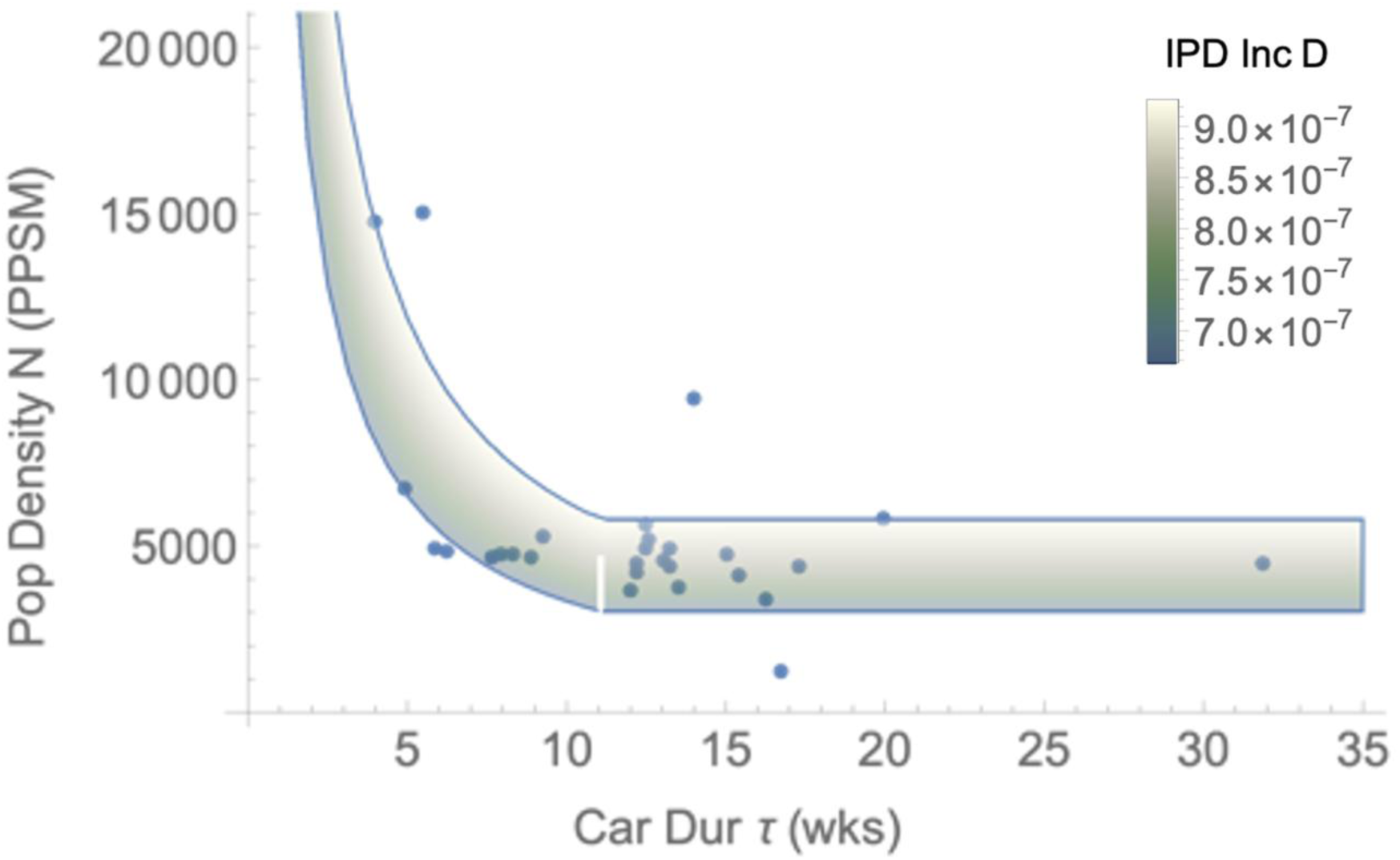
Fitting population density and invasive incidence using a piecewise contact rate function indicates Spn transmission is density dependent. Spn serotype average population density data (blue dots) and the model fit (green-to-white gradient, 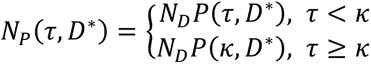 , where 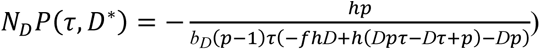 of population density as a function of carriage duration τ and equilibrium invasive incidence *D\**. Model fits support the hypothesis Spn transmission is density-dependent and that shorter-duration serotypes are more likely to cause invasive disease in higher-density areas. The model fitting produced the parameter estimates *b_D_* = 6.67x10-5, *κ* = 11.07, and *f_P_* = 229. Additional fixed parameter values are invasive clearance *h* = 0.5 and probability of invasion *p* = 0.00029.

### Sensitivity Analysis

While both the power and piecewise contact rate models show that Spn transmission is density-dependent, they differ in the strength of this dependence. The power contact rate model suggests nearly pure (*r =* 1) density-dependence while the piecewise model supports a hybrid transmission function that plateaus at higher densities. Each model has several parameters controlling the level of density-dependence that we fitted in this analysis. We now assess how robust these model results are to varying these parameters by carrying out two sensitivity analyses. First, to get an overall sense of parameter sensitivity we generated a parameter combination plot (Figure 6A, C) by sampling 500 datapoints for each estimated parameter from the distributions listed in methods Table 3. Each parameter combination was plotted as a grey line and all 500 plots were overlayed with the empirical observations in blue. The monotonically decreasing relationship between carriage duration and population density holds for all 500 curves for both the power contact rate model (Figure 6A) and the piecewise contact rate model (Figure 6C).

**Figure 6:**
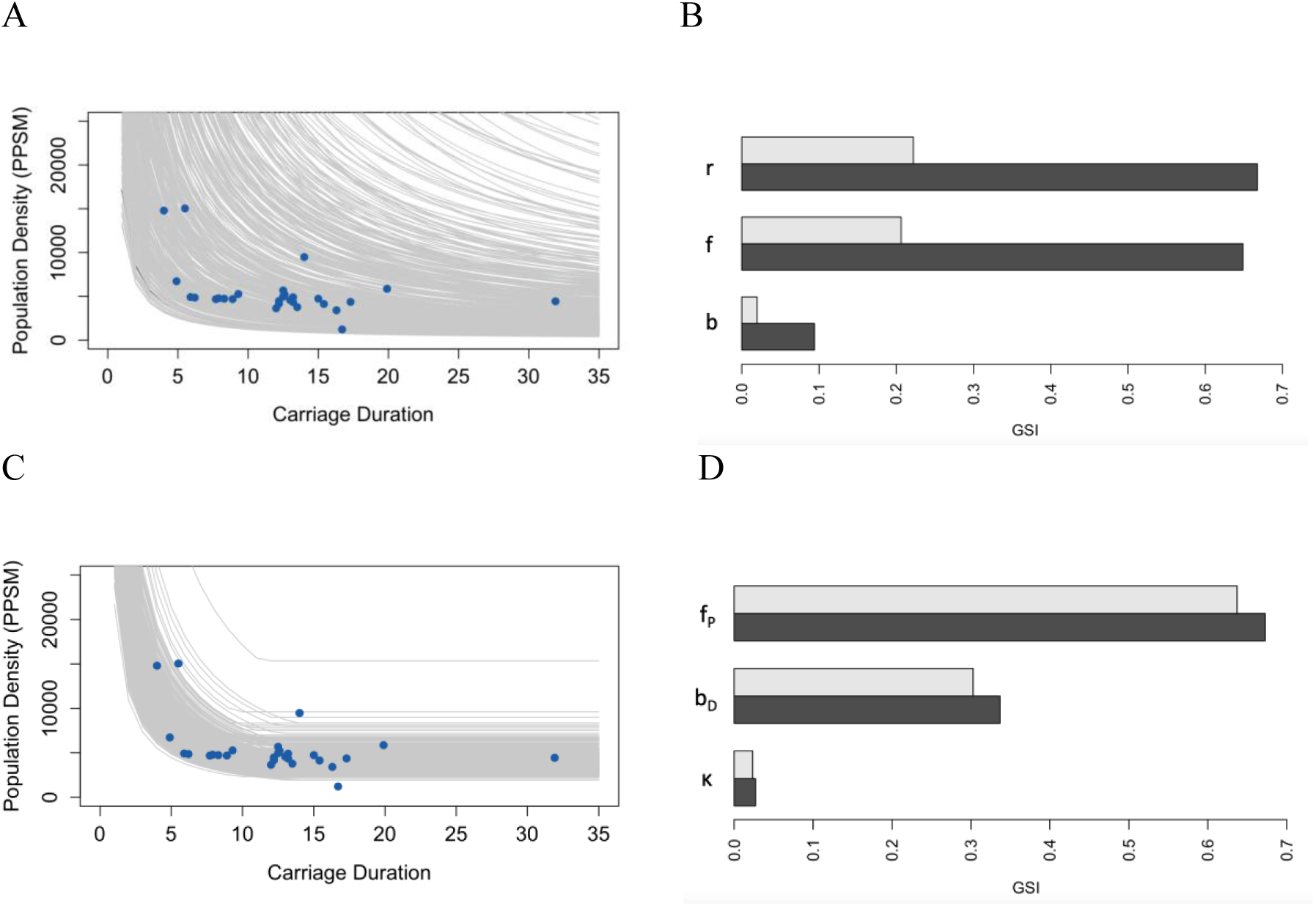
Sensitivity analysis of the population density functions (equations 4 and 9) reveals that the model fitting results are robust across a wide range of parameter values. A) and C) Parameter combination plots of equations 4 (Figure 6A) and 9 (Figure 6C) (see methods) generated by sampling 500 sets of parameter estimates from the distributions described in Table 3 (grey lines) overlayed with Spn serotype average population density data (blue dots). Both graphs show that even with wide variation in parameter values the overall negative association between carriage duration and population density generally holds. B) First order (grey bars) and total (black bars) generalized sensitivity indices (GSI) for the population density function based on the power contact rate model (equation 4). Results show that population density is mainly influenced by changes in the power exponent parameter *r* and immune protection parameter *f* with a smaller contribution from the transmission rate constant *b*. D) First order (grey bars) and total (black bars) generalized sensitivity indices (GSI) for the population density function based on the piecewise contact rate model (equation 9). The model was most sensitive to changes in the immune protection parameter *f_P_* and the density-dependent transmission constant *b_D_*.

**Table 2:** Definitions for the variables and parameters used in the compartmental epidemiological model along with values and reference sources used in the model fitting.

| **Variables** |  |  |
| --- | --- | --- |
| **Definition (Units)** |  |  |
| $S(t)$ | Proportion of individuals in the susceptible class (at risk of acquiring the focal Spn serotype) at time $t$ | |
| $C(t)$ | Proportion of individuals in the carriage class (carrying the focal Spn serotype) at time $t$ | |
| $I(t)$ | Proportion of individuals in the invasive class (with an invasive infection caused by the focal serotype) at time $t$ | |
| $R(t)$ | Proportion of individuals in the recovered class (individuals who have cleared an infection and whose immunity offers protection from reacquiring the same serotype) at time $t$ | |
| **Fixed Parameters from Prior Literature** |  | **Value**<br>**(Reference)** |
|  | **Definition (Units)** |  |
| $p$ | Probability of progressing to the invasive state at the time of carriage acquisition | 0.00029<br>(Metcalf et al., 2023) |
| $\tau$ | Average duration of carriage for a given Spn serotype (weeks) | Serotype dependent<br>(Sleeman et al., 2006) |
| $h$ | Rate of transition from the invasive to recovered class, due to pathogen clearance (per week) | 0.5<br>(Baldo et al., 2015) |
| $N$ | Population density (persons per square mile) | Serotype dependent<br>(Langley et al., 2015) |
| **Estimated Parameters** |  |  |
|  | **Definition (Units)** |  |
| $f$ | Power contact rate model duration of immune protection (weeks) | |
| $r$ | Power contact rate function exponent | |
| $b$ | Power contact rate model transmission rate constant (per person per week) | |
| $\kappa$ | Carriage duration threshold constant for the piecewise contact rate model (weeks) | |
| $b_D$ | Piecewise contact rate model density-dependent transmission rate constant | |
| $f_P$ | Piecewise contact rate model duration of immune protection (weeks) | |

**Table 3:** The distribution function and sampling boundaries used in the GSI sensitivity analyses testing the inverse relationship between population density and carriage duration. Parameter ranges are modified from Table 1 to use truncated Normal distributions where required to avoid parameter selections outside of biological limits (for parameters *r* and *b*). Parameter combination plots are obtained by sampling from the normal and truncated normal distributions to generate 500 sets of parameter estimates for both the power and piecewise contact rate models. When designating the lower and upper boundary constraints for the GSI factorial design, we use the 2.5^th^ and 97.5^th^ percentiles representing 95% of the normal or truncated normal probability distributions respectively.

| **Power Law Contact Rate Model** |  |  |  |  |
| --- | --- | --- | --- | --- |
| **Parameter** | **Distribution** | **Arguments** | **Lower Bound** | **Upper Bound** |
| $b$ | Trunc Normal | Mean = 7.69e-5<br>SD = 8.32e-5 | 6.35e-6 | 2.47e-4 |
| $f$ | Normal | Mean = 237<br>SD = 23.4 | 191 | 282 |
| $r$ | Trunc Normal | Mean = 0.988<br>SD = 0.139 | 0.680 | 0.995 |
| **Piecewise Contact Rate Model** |  |  |  |  |
| **Parameter** | **Distribution** | **Arguments** | **Lower Bound** | **Upper Bound** |
| $f_p$ | Normal | Mean = 229<br>SD = 19.8 | 190 | 268 |
| $b_D$ | Normal | Mean = 6.67e-5<br>SD = 1.28e-05 | 4.16e-05 | 9.18e-05 |
| $\kappa$ | Normal | Mean = 11.07<br>SD = 1.27 | 8.58 | 13.56 |

We next evaluate the relative impact of each parameter and whether their effects are stable across carriage durations. To do this we perform a sequential sensitivity analysis using the ‘multisensi’ R package with carriage duration acting as the sequential variable. The parameter ranges used are the 95% confidence interval ranges listed in methods Table 3. For the power contact rate model, Figure 6B shows the generalized sensitivity indices (GSI) for each parameter and reveals that the power contact rate function exponent *r* and the immune duration parameter *f* had the most impact on the model with the *b* parameter having relatively less influence. A graph of the variation in sensitivity indices across carriage duration indicates that the main effects are stable across serotype durations (Figure S4A). The piecewise contact rate model was most sensitive to the *f_P_* and *b_D_* parameters and least impacted by the threshold parameter *κ* (Figure 6D). However, looking at the variation of the indices across carriage duration (Figure S4B) shows the impact of *κ* increases when the model transitions from density- to frequency-dependent transmission.

Lastly, our epidemiological model assumes that the risk of progressing into invasive disease (parameter *p*) is constant. We fixed *p* to 0.00029 based on a previous model fit (Metcalf et al., 2023). However, it has been shown that invasiveness, as estimated by invasive attack rate, varies considerably across serotypes (Sleeman et al., 2006). To assess the robustness of our model across a range of *p*, we refit the piecewise model using values from the 95% confidence interval derived from our earlier model. We show in Figure S5, that even with highly divergent values of *p*, the model performs similarly with comparable estimates of 𝜅, *b_D_*, and *f_P_* (Table S2).

## Discussion

In this investigation we examined the role of human population density in Spn transmission and tested the hypothesis that shorter carriage duration serotypes are attenuated in lower density areas, given they have less time to successfully transmit to a new host as compared to their longer carriage duration counterparts. In agreement with our hypothesis, we show that (i) Spn serotype distributions are structured by host population density (Figure 2), (ii) human density correlates with serotype carriage duration and attack rate (Figures 3, 5), and (iii) serotype transmission is both frequency- and density-dependent (Figure 4). Together these results show that serotypes are differentially impacted by human population density, with short carriage duration serotypes less likely to transmit (and cause invasive disease) in less populated areas.

These results have important and generally overlooked implications for pneumococcal epidemiology, given the substantial variation in human population density around the world, and the connections between carriage duration and other traits. Carriage duration is a key life-history trait that impacts many other aspects of Spn biology including antimicrobial resistance, recombination, invasiveness, and serotype prevalence (Chaguza et al., 2016, Kalizang’oma et al., 2021, Sleeman et al., 2006, Li et al., 2013). For example, longer carriage duration serotypes tend to have higher carriage prevalence within a community (Li et al., 2013, Weinberger et al., 2009, Högberg et al., 2007), although these prior studies did not unpack the role of variation in human population density. Our results indicate this advantage can be attenuated in regions with high population densities that allow greater transmission of shorter carriage duration serotypes.

Our results imply that pneumococcal surveillance systems that operate across different distributions of population densities may report different distributions of serotypes even if the underlying serotype diversity is the same. This may play a role in the observed geographical variation of IPD incidence caused by non-vaccine serotypes following the introduction of pneumococcal conjugate vaccine (Lewnard and Hanage, 2019). Also, previous work has identified a positive correlation between beta-lactam resistance and carriage duration, likely due to increased antibiotic exposure in strains with longer human residency times and increased opportunity for recombination with beta-lactam resistant *Streptococcus mitis* (Lehtinen et al., 2017, Kalizang’oma et al., 2021). Our investigation would therefore suggest that beta-lactam resistance may be more likely to occur in less populated areas. An important area of future work is to determine if beta-lactam resistance is inversely correlated to population density and to tease apart how beta-lactam antibiotic prescription rates influence this potential relationship.

More broadly, our results offer a route-map for investigation of density-dependent transmission in other human pathogens with complex carriage versus invasive epidemiological dynamics, as seen in various opportunistic bacterial pathogens (e.g. meningococcus, group A *Streptococcus*). The introduction of density-dependence to bacterial epidemiological models also raises new opportunities to revisit classical epidemiological questions in light of density-dependent effects, including critical community sizes (Finkenstädt et al., 1998, Bartlett, 1960), vaccine thresholds (Anderson and May, 1991), and the evolution of virulence (Alizon et al., 2009).

This study has several limitations. The average census tract population density for the ABCs catchment area (4600 PPSM) is considerably higher than the average population density of the U.S. population (94 PPSM; (Bureau, 2021)), reflecting a surveillance focus on more densely-populated regions of the country. Our results indicate that this surveillance choice may bias the distributions of serotypes (and potentially other pathogens) that are reported. Also, census tract information for our surveillance dataset was missing for 1,506 of 13,798 case-patients (11% of total). For parameter estimation, the quality of the model fits varied widely depending on the algorithms and arguments used. We evaluated several global and local optimization procedures (e.g. NMinimize, Newton, InteriorPoint) using the corrected Akaike Information Criterion (AICc), but our comparisons were not exhaustive and better parameter estimations may be possible. The Spn average carriage duration and invasive attack rate data used in this analysis are the most comprehensive available but were acquired more than 15 years ago as part of a series of longitudinal pediatric studies in the United Kingdom (Sleeman et al., 2006) and may not capture unique features of current adult invasive disease epidemiology (Alanee et al., 2007). Also, the carriage duration, invasive attack rate, invasive incidence, and host population density data are averaged by serotype which does not account for within-serotype variation or measurement error inherent within these estimates.

Our analysis shows that more invasive short carriage duration serotypes spread more easily in communities with higher population densities (Figure 3). Importantly, in regions where population or housing density is associated with lower community socioeconomic status, these shorter duration, higher invasive serotypes may be preferentially affecting the very communities that are least equipped to intervene or respond. A focus for follow-up work will be to examine the implications of density-dependent pneumococcal transmission in the context of the social determinants of health.

## Methods

### Study Collection

The CDC Active Bacterial Core surveillance (ABCs) program conducts active laboratory and population-based surveillance for invasive Spn infections (in 10 states (complete states or selected counties), representing more than 34.4 million persons (https://www.cdc.gov/abcs/reports-findings/survreports/gas18.html). ABCs defines an invasive Spn case as illness in a surveillance area resident with isolation of Spn from a normally sterile site. Surveillance staff at sites complete a standardized case report form that includes basic demographic characteristics and risk factors of infection on all cases, and coordinate collection and transfer of isolates to CDC’s *Streptococcus* laboratory for characterization. In total, 11,245 available invasive Spn isolates from cases identified in 2016-2019 were included in the analysis.

We calculated the average population density for serotypes acquired from invasive disease cases that were within the catchment area of the ABC surveillance program from 2016 through 2019. We used data obtained from 2015-2019 5-Year estimates of the American Community Survey (ACS) and U.S. Census Bureau’s 2021 Planning Database (PDB) with both census tracts and ZIP Code Tabulation Areas (ZCTAs) as the geographic units of measure. To calculate the average population density for each Spn serotype acquired through ABC surveillance at the ZCTA level, we used ZCTAs obtained from the 2015- 2019 American Community Survey 5-Year Estimates dataset B01003 to infer population size. Geographic area for each ZCTA was obtained using the 2017 land area data from the Gazetteer file ’ZIP Code Tabulation Areas’. For census tract population density measurements, we used the U.S. Census Bureau’s 2021 Planning Database (PDB) which provides both population count and land area data at the census tract level.

Population density for each census tract or ZCTA was calculated by dividing the population size by its land area. A population density was associated with each invasive case by linking the ZCTA or census tract population density with the zip code or census tract recorded in the case report form. The average host population density for each serotype was calculated by averaging the census tract or ZCTA population densities associated with each invasive pneumococcal disease case caused by that serotype. To ensure the ZCTAs correspond to the correct zip code we validated each pairing using the crosswalk database created by the Census Reporter project (https://censusreporter.org/). Serotype invasive incidence was obtained by dividing serotype IPD case counts by total serotype population size. Serotype population size was calculated by identifying the unique set of census tracts where an IPD case with that serotype was found and summing their individual population sizes.

Carriage duration data was obtained from a study from (Sleeman et al., 2006). We assumed that the carriage duration for serotype 4 was too short to be reliably detected given the sampling times of the longitudinal study, so we imputed a conservative estimate of four weeks (the longest sampling interval used in Sleeman *et al*.) instead.

### Epidemiological Model Description

#### Power Contact Rate Model

To analyze how carriage duration interacts with population density to effect serotype transmission, we construct a compartmental epidemiological model of pneumococcal transmission based on previous work (Metcalf et al., 2023). In this model framework, host individuals are classified as being either susceptible, infected or recovered and immune, with regard to a specific, focal serotype. The infected class is further broken down into a carrier state and an invasive state. The proportions of individuals in a susceptible, carrier, diseased and recovered class for a focal strain are denoted by the variables *S*, *C*, *I* and *R* respectively (Figure 7), and their dynamics are given by the following system of four ordinary differential equations where the force of infection *λ* = *bN^r^C*

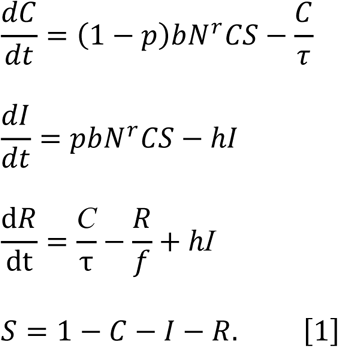

**Figure 7:**
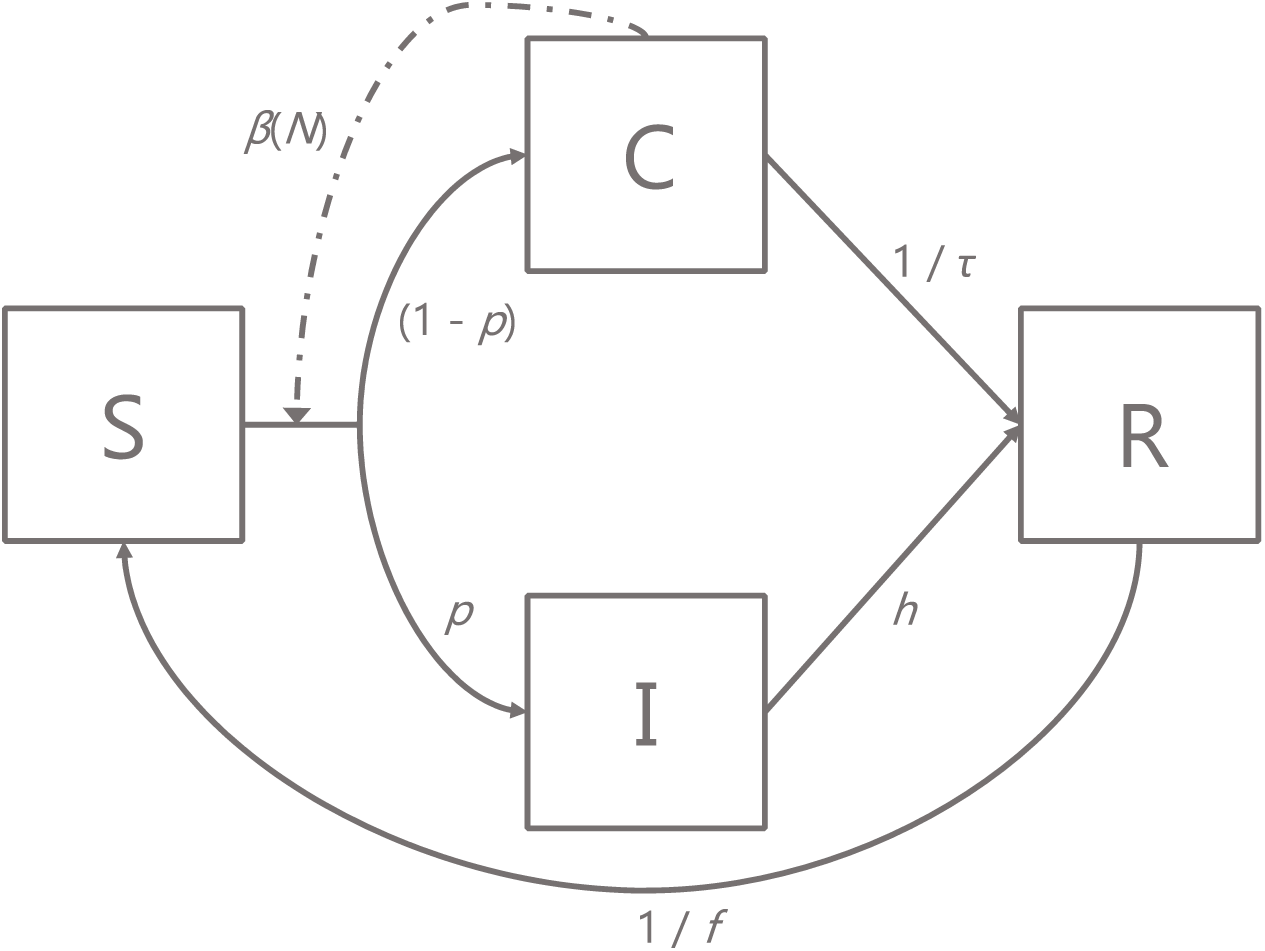
Schematic diagram of the epidemiological model. Boxes represent proportions of hosts in mutually exclusive states: susceptible (*S*), infected asymptomatic carriers (*C*), invasive (*I*) or recovered and immune (*R*). Solid arrows represent flows of individuals between states, and dashed arrows represent factors influencing those flows. Equations describing the system are presented in the main text (methods equations 1), along with parameter definitions (Table 2). See Metcalf et al. for initial model development (Metcalf et al., 2023).

Recall that the force of infection *r* parameter specifies how population density impacts transmission (Fig 1). Depending on the value of *r*, the power contact density function allows for a purely density-dependent (*r*=1) or frequency-dependent (*r*=0) host contact structure (Borremans et al., 2017). An *r* value between 0 and 1 generates a hybrid contact density function that has an intermediate saturating shape which is more strongly density-dependent at lower population densities but plateaus at higher densities (Borremans et al., 2017). The model assumes transmission is from carriers only, there are no multiple infections, and that intrinsic virulence, the core set of traits that concern adhesion, invasion, and proliferation in disease sites, are equivalent for all serotypes. In other words, the invasive disease parameter *p* is held constant across serotypes with different carriage durations. Finally, all rates are defined using a time unit of one week. The model variables and parameter definitions are detailed in Table 2.

Solving the system of differential equations representing the SCIR model at a stable endemic equilibrium [*S*\*, *C*\*, *I*\*, *R*\*] yields the following,

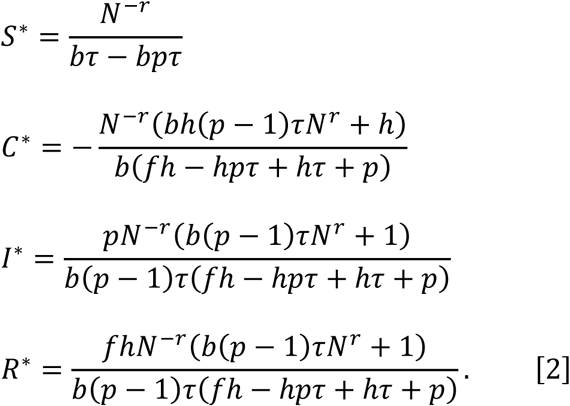

A key aim for this analysis is to estimate the level of density-dependence within the host contact structure as represented by the power contact rate function exponent *r*, using serotype-specific epidemiological data on carriage duration 𝜏, population density *N*, and invasive incidence *D*. To do this we first derive the steady state invasive incidence *D\** from the equilibrium solutions to this system of differential equations by defining a function based on new invasive cases (*D* =* 𝑝𝑏𝑁^𝑟^𝐶^∗^𝑆^∗^) yielding,

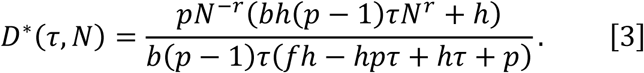

We then derive an expression for population density as a function of carriage duration and invasive disease abundance by rewriting the equation for invasive incidence at equilibrium (*D*\* equation 3 above) in terms of population density *N*,

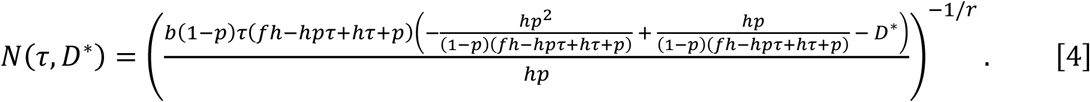

#### Piecewise Contact Rate Model

We also present an alternative epidemiological compartmental model that employs a piecewise transmission term function with a defined density-dependent and frequency-dependent component. Besides the transmission term, the overall structure of the model is unchanged from Figure 7. The model dynamics are given by a system of four ordinary differential equations,

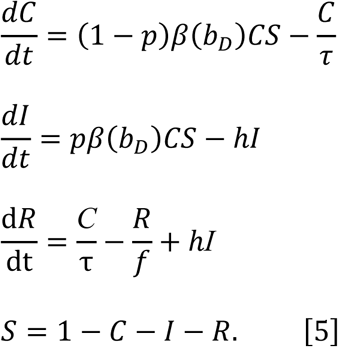

Where the transmission term 𝛽(𝑏_𝐷_) is conditional on the threshold carriage duration parameter 𝜅,

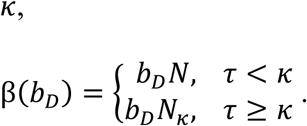

The threshold carriage duration parameter 𝜅 marks the transition point from density- to frequency-dependent transmission. This piecewise threshold cutoff would naturally be defined in terms of population density instead of carriage duration. It is required to be written in terms of carriage duration here as a constraint of fitting two functions, population density and invasive incidence, simultaneously. At least one parameter, in this case carriage duration, needs to link the two functions together to use both equations to fit shared parameters. However, once we estimate 𝜅 we can plug that value into our function for population density *N* (equation 8, derived below) to represent the cutoff threshold in terms of *N* (parameter 𝑁_𝜅_). At population density 𝑁_𝜅_, the transmission coefficient *β* transitions to frequency-dependence where β is fixed at the threshold value (𝑏_𝐷_𝑁_𝜅_) and invariant across higher population densities (Figure 4). The endemic equilibrium [*S*\*, *C*\*, *I*\*, *R*\*] solutions for the piecewise contact rate model are provided below,

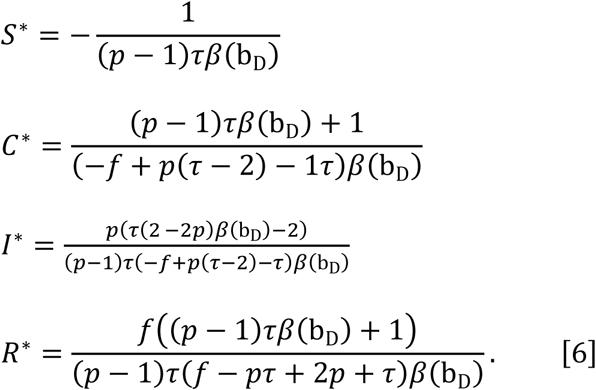

Similar to the power contact rate model, the piecewise model steady state invasive incidence *D_p_\** is based on new invasive cases from the density-dependent (*D_D_* =* 𝑝𝑏_𝐷_𝑁𝐶^∗^𝑆^∗^) and frequency-dependent (*D_F_* =* 𝑝𝑏_𝐷_𝑁_𝜅_𝐶^∗^𝑆^∗^) components yielding,

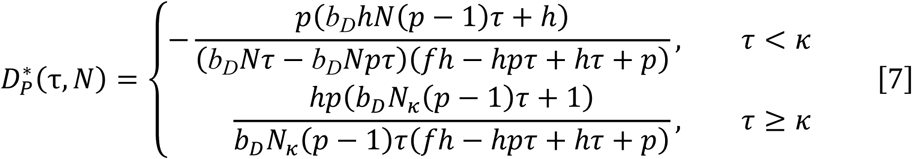

We next derive a piecewise expression of population density as a function of carriage duration τ and invasive incidence *D*. For the density-dependent component, we rewrite the equation for invasive incidence at equilibrium (*D_D_*\*) in terms of population density *N*,

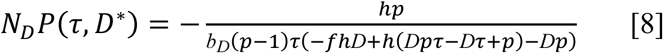

By definition, we cannot rewrite the frequency-dependent invasive incidence equation in terms of *N* since it is not a function of *N*. Instead, we specify that past the piecewise threshold cutoff carriage duration *κ*, serotypes will transition from density- to frequency-dependent transmission, and both the contact and transmission rates will hold constant at that level across all larger population densities. Thus, the piecewise function for population density will be the following,

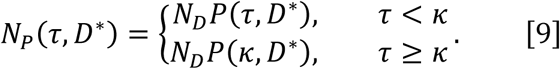

### Statistical Analysis

We use the ‘NonlinearModelFit’ function from Mathematica 11.1.1.0 to perform model fitting for both the power and piecewise contact rate models. We simultaneously fit the population density and invasive incidence data captured by ABCs by defining equations for population density and invasive incidence as components of a piecewise function. Parameters are then estimated for both the power contact rate model (equations 3 and 4) and the piecewise contact rate model (equations 7 and 9). The data used for model fitting is provided in Table S1. For the power contact rate model, we use the Automatic method which enables Mathematica to choose a model fitting algorithm based on the data and model. For the piecewise contact rate model, we use the NMinimize global optimization algorithm employing the Differential Evolution function minimizer with the parameters CrossProbability = 0.1 and ScalingFactor = 0.9. We also specify that local search be performed by the QuasiNewton method using the PostProcess argument. A sensitivity analysis of the model fit employing equations 4 and 9 is performed using the ‘multisensi’ R package v2.1-1. For the power contact rate model, parameter ranges are calculated using the 95^th^ percentiles of either the Normal distribution (parameter *f*) or the truncated Normal distribution (parameters *r* and *b*). The truncated Normal distribution is required to prevent negative *b* values and values of *r* that exceeded the limit case (*r* = 1) (Table 3). Ranges for the *f_P,_ b_D,_* and *κ* parameters of the piecewise contact rate model are all derived from the Normal distribution (Table 3). The Normal and truncated Normal mean and standard deviation values come from the best fit model estimates. The truncated normal distributions are generated using the ‘truncnorm’ R package v1.0-9. All additional statistical analyses are carried out using the base package of R v4.3.3 and all plots are created using either the R base package, the R ‘ggplot2’ v3.5.0 package, or Mathematica 11.1.1.0. Only serotypes with at least 5 invasive cases were included in the analysis.

## Disclaimer

The findings and conclusions in this article are those of the authors and do not necessarily represent the official position of the CDC.

This activity was reviewed by the CDC Institutional Review Board, deemed not research, and was conducted consistent with applicable federal law and CDC policy (see e.g., 45 C.F.R. part 46.102(l)(2), 21 C.F.R. part 56; 42 U.S.C. §241(d); 5 U.S.C. §552a; 44 U.S.C. §3501 et seq.).

## Supplementary

Epidemiological compartmental models require that a contact structure be specified. These contact structures are usually frequency-dependent or density-dependent. We performed a literature survey to estimate how much each contact structure is used in the field.

The first step in this analysis is to identify the papers that researchers use when citing density-dependent contact structures for their work. To accomplish this, we used the Publish or Perish tool (https://harzing.com/resources/publish-or-perish) to search for selected keywords associated with density-dependent compartmental models in Google Scholar and sort the matching papers by year. We generated a collection of 148 pneumococcal articles using the following keyword search terms:

> "density dependence"|"density dependent" -"cell density" transmission susceptible "streptococcus pneumoniae"|pneumococcus|pneumococcal "compartmental model"|"epidemiological model"|"dynamic model"|"mathematical model"|"transmission model"

Using the R statistical programming language, we then filtered out all papers that didn’t contain pneumoniae, pneumococcal, pneumococcus or PCV (pneumococcal conjugate vaccine) in the title:

setwd(’/path/Spn_contact-struc_litReview_1-1-2023/’)
df1 <- read.delim("Spn_contact-dens_Rev_PoP_qry-GQ6_1-5-2023.csv",header=TRUE,sep=’,’, fill=TRUE)
df2 <- df1[grepl("pneumoniae|pneumococcal|pneumococcus|pcv", tolower(df1$Title)),]

This left 5 remaining articles:

1. "Pneumococcal capsule blocks protection by immunization with conserved surface proteins"
2. "Frequency-dependent selection in vaccine-associated pneumococcal population dynamics"
3. "Periodicity of epidemics of invasive disease due to infection with Streptococcus pneumoniae in the United States"
4. "Estimating the transmission parameters of pneumococcal carriage in households"
5. "Estimating the transmission parameters of pneumococcal"

Papers 1-3 didn’t refer to density dependence within the context of epidemiological modeling and were removed. Papers 4 and 5 refer to the same study and is the only article to incorporate density dependent transmission although it was not at the population level. Their work focused on how household size impacted Spn spread. Finally, to validate the google scholar results, we performed a more comprehensive search of *Streptococcus pneumoniae* modeling papers using the PubMed citation database. We generated a collection of 100 pneumococcal articles using the following search terms:

> (("streptococcus pneumoniae"[Title/Abstract]|pneumococcus[Title/Abstract]|pneumococcal[Title/Abstract]) AND ("compartmental model"[Title/Abstract]|"epidemiological model"[Title/Abstract]|"dynamic model"[Title/Abstract]|"mathematical model"[Title/Abstract]|"transmission model"[Title/Abstract]))

We then searched the 50 most recent papers for “frequency dependence|dependent” or “density dependence|dependent” keywords. We found no additional pneumococcal transmission papers that were employing a density dependent contact function.

## Data Availability

All data produced in the present work that are not considered personally identifiable information are contained in the manuscript

---

**Figure S1:**
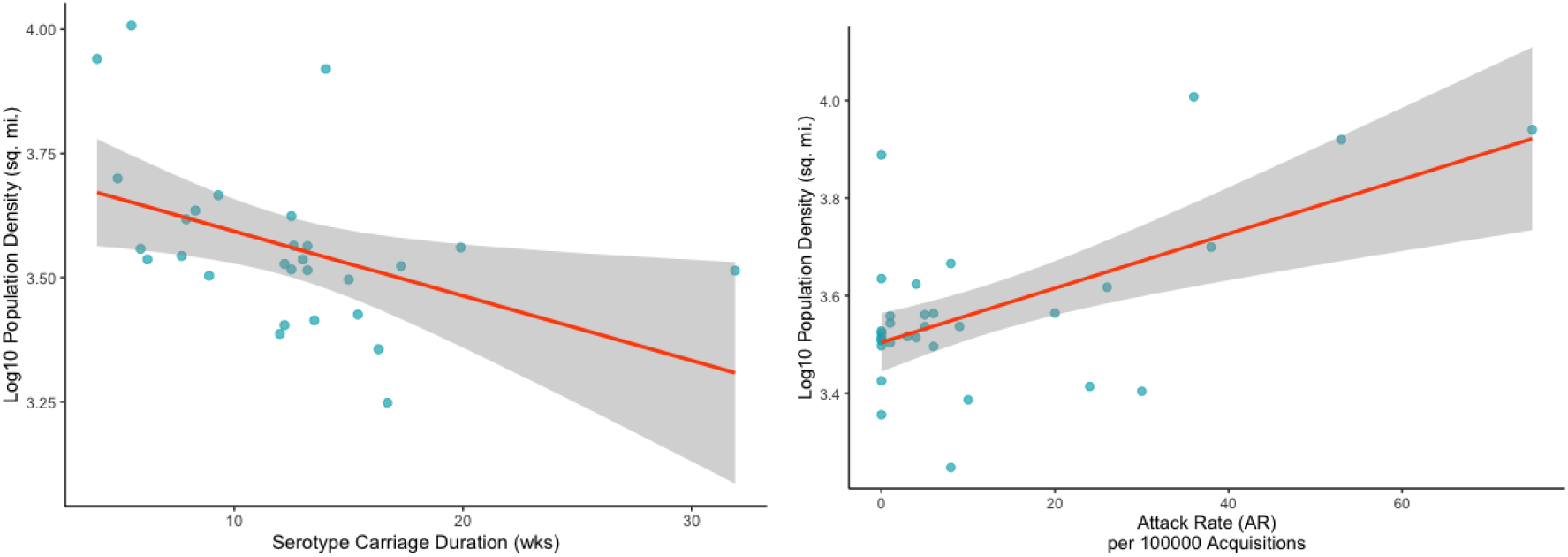
Average serotype population density has a negative correlation with carriage duration and a positive correlation with invasive attack rate (AR). (A) Serotype carriage duration (blue dots) against log10 average serotype population density (measured using ZCTAs) yields a significant inverse association (Kendall Tau = -0.385, p-value = 0.00415). (B) Invasive AR (blue dots) against log10 average serotype population density (measured using ZCTAs) yields a significant positive association (Kendall Tau = 0.256, p-value = 0.048). These relationships indicate that shorter duration and more highly invasive serotypes are more likely to cause IPD in denser environments.

**Figure S2:**
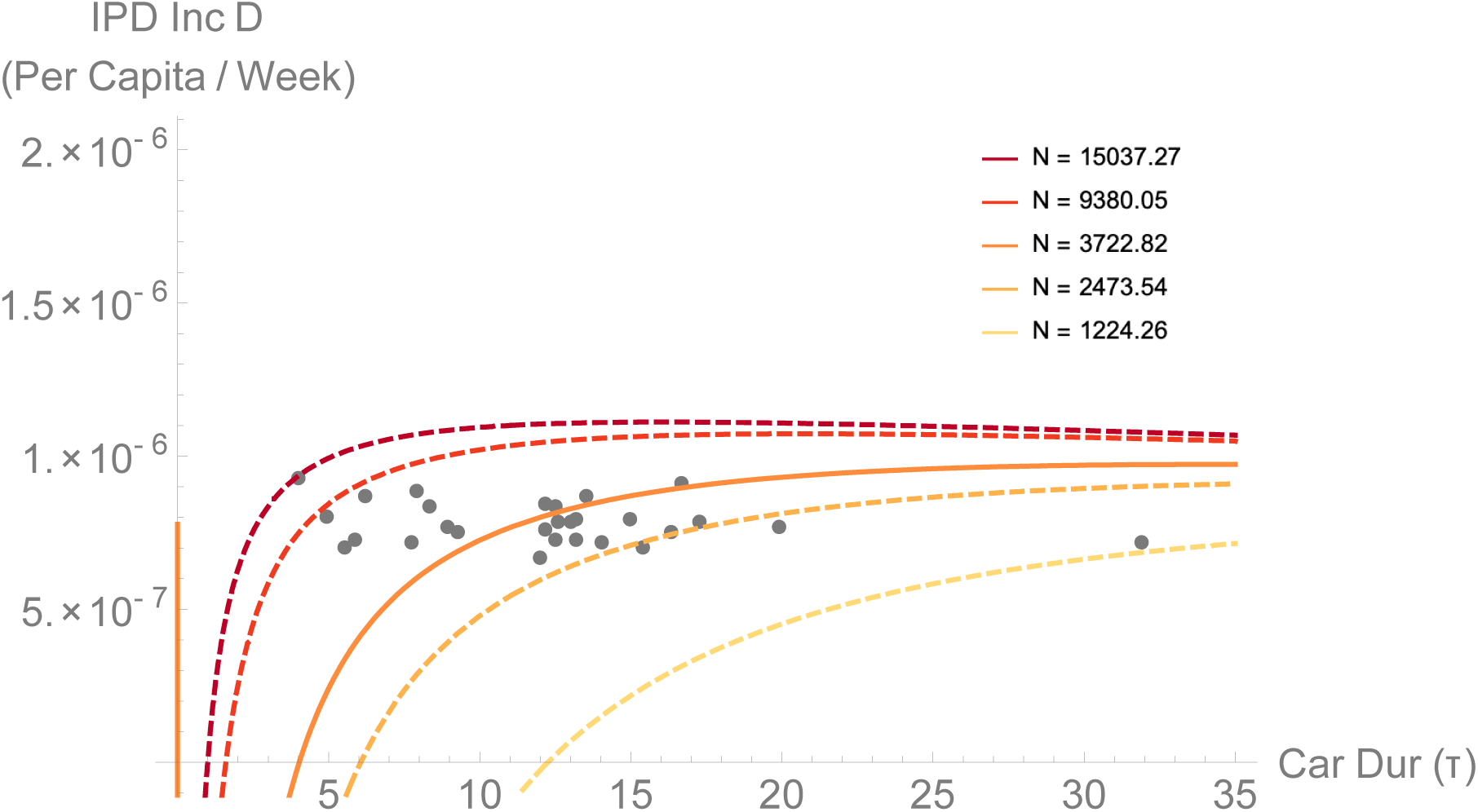
Fitting invasive incidence using a piecewise contact rate function provides support for Spn density dependent transmission. Spn per capita invasive incidence data in weeks (black dots) and the model fits (red-to-yellow gradient lines, 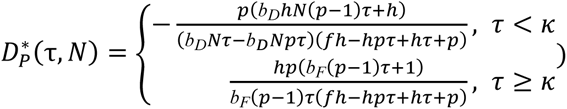 of invasive incidence as a function of carriage duration *τ* and population density *N*. The model fits support the hypothesis that Spn transmission is density-dependent and that shorter-duration serotypes are more likely to cause invasive disease in higher-density areas. Population density *N* was set to 𝑁_𝜅_ for the range of population densities observed in the data (1224.26 PPSM to 15037.27 PPSM), making the density- and frequency-dependent transmission coefficients equivalent yielding a set of continuous curves across the two piecewise components. The solid line, 𝑁_𝜅_ = 3722.82 PPSM, represents the best-fit line and the black vertical line designates the piecewise threshold constant *κ* = 11.07.

**Figure S3:**
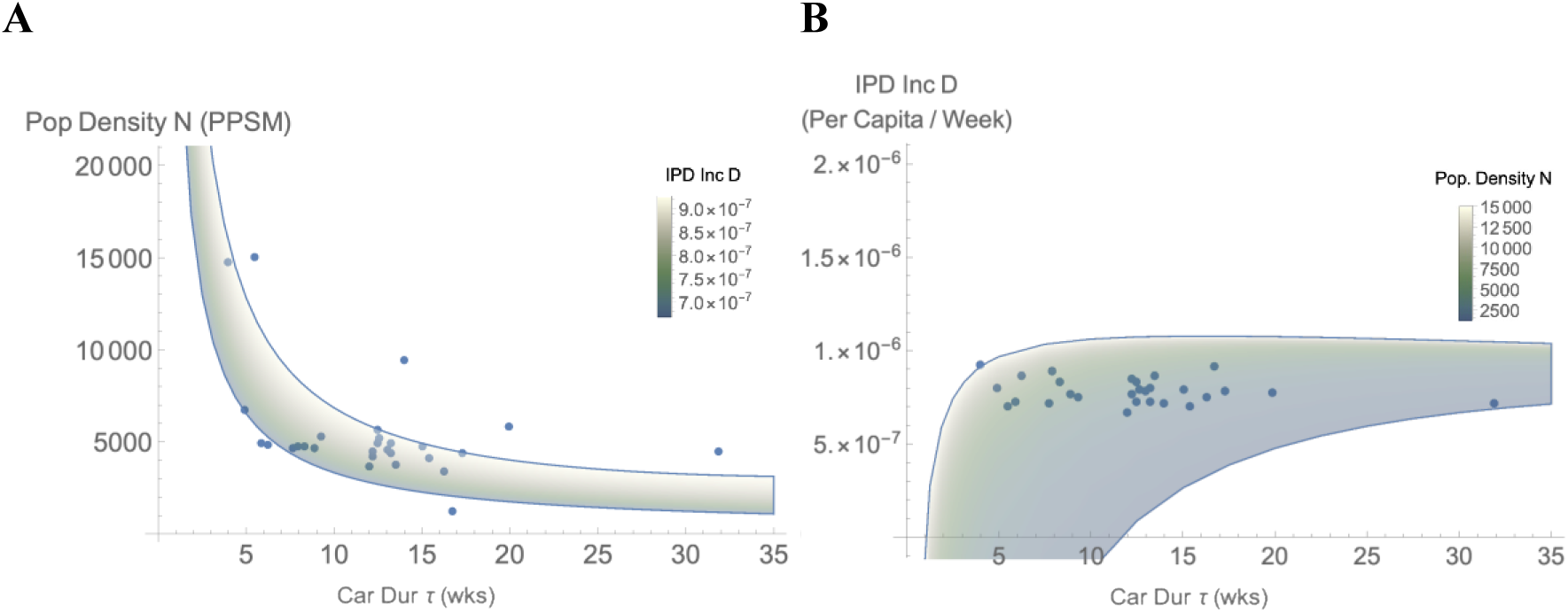
Fitting population density and invasive incidence using a power contact rate function indicates Spn transmission is density dependent. A) Spn serotype average population density data (blue dots) and the model fit (green-to-white gradient, 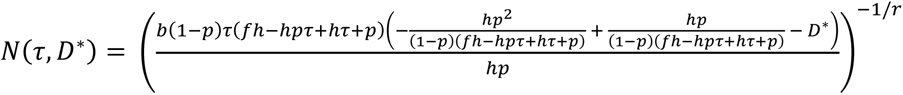 of population density as a function of carriage duration τ and equilibrium invasive incidence *D\**. B) Spn per capita invasive incidence data in weeks (blue dots) and the model fit (green-to-white gradient, 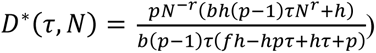 of invasive incidence as a function of carriage duration *τ* and population density *N*. Both model fits support the hypothesis Spn transmission is density-dependent and that shorter-duration serotypes are more likely to cause invasive disease in higher-density areas. The model fitting estimated the power contact rate exponent *r* = 0.988 (i.e., Spn transmission is constrained by a nearly linear contact density function), the immune duration parameter *f* = 237, and the transmission rate constant parameter *b* = 0.0000769. Additional fixed parameter values are invasive clearance *h* = 0.5 and probability of invasion *p* = 0.00029.

**Figure S4:**
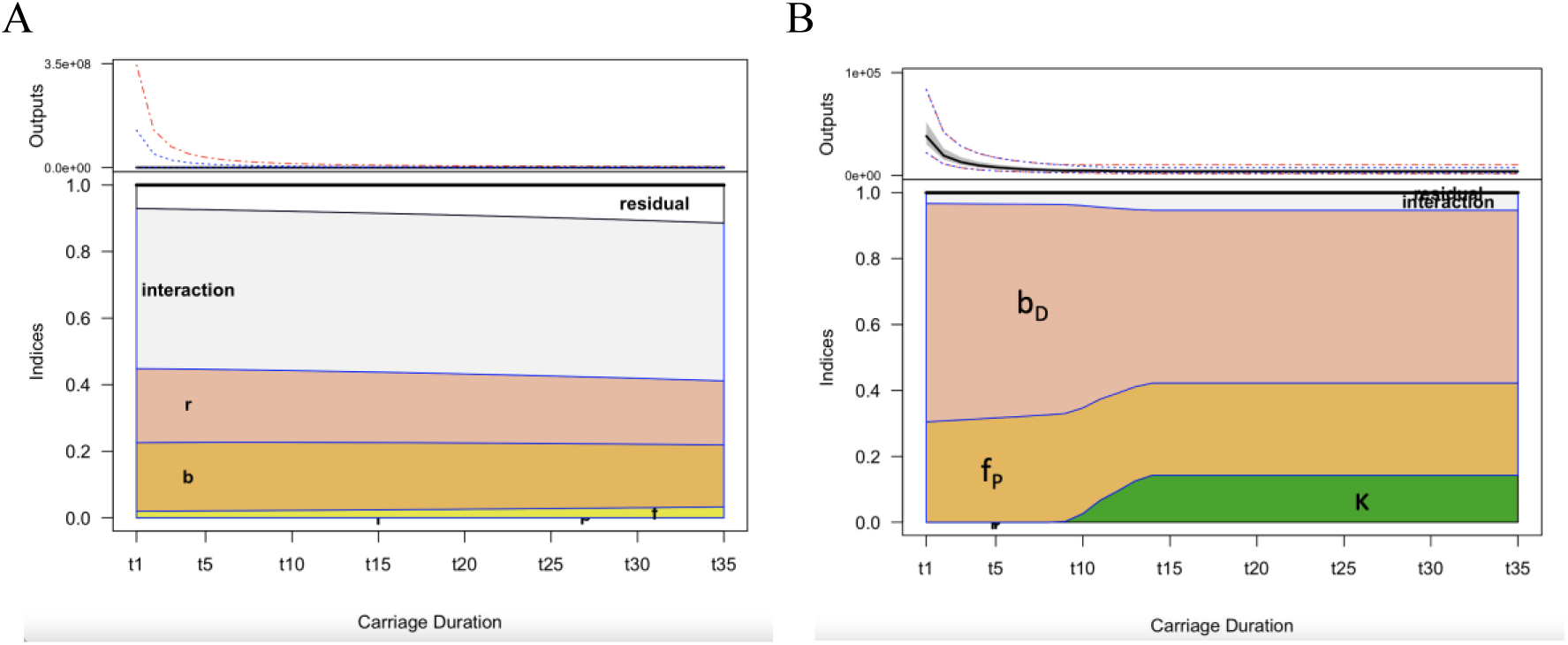
**Additional Sensitivity plots of the population density functions (equations 4 and 9).** A) Plot of the sensitivity indices of the power contact rate density function (equation 4) from *τ* = 1 to 35 reveals that the sensitivity of population density to the main effects of *r, f, and b* remain stable across the range of carriage durations. B) Plot of the sensitivity indices of the piecewise contact rate density function (equation 9) from *τ* = 1 to 35 indicates the *f_P_* and *b_D_* parameters remain stable across carriage durations. Sensitivity to the threshold parameter *κ*, however, increases as the model transitions from density- to frequency-dependent transmission.

**Figure S5:**
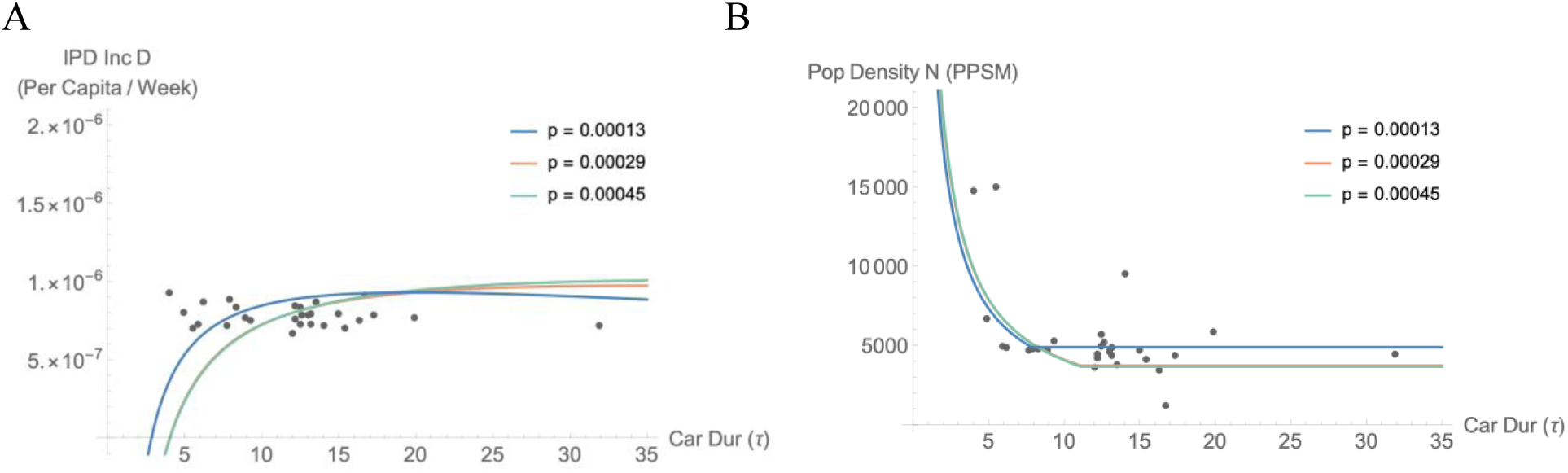
The relationship between population density and carriage duration in the piecewise model holds across wide variations in invasiveness. The graphs represent population density (panel A) and invasive incidence (panel B) as functions of carriage duration fitted with the invasiveness parameter *p* set to 0.00013 (blue curve), the baseline 0.00029 (red curve), and 0.00045 (green curve). The values of parameter *p* represent estimates at the 2.5th, median, and 97.5th percentiles generated from a previous model fit (Metcalf et al., 2023). All three curves on each plot show strong concordance indicating that model performance is maintained across a wide range of invasive risk.

**Table S1:** Pneumococcal life-history traits and host population metadata aggregated by serotype. This set of observations was created by combining the longitudinal data from Sleeman et al. with population density data from United States Census Bureau’s 2021 Planning Database. This table provides invasive case counts (IPD case count), locations sampled (State Count), census tracts sampled (Census Tract Count), average census tract population density (Avg. Pop. Dens.), total population size (Tot. Pop. Size), per capita IPD rate (IPD Rate), and carriage duration.

| **Spn Serotype** | **IPD Case Count** | **State Count** | **Census Tract Count** | **Avg. Pop. Dens. (sq. mi.)** | **Tot. Pop. Size** | **IPD Rate (per capita)** | **Avg. Carriage Duration (weeks)** |
| --- | --- | --- | --- | --- | --- | --- | --- |
| 4 | 183 | 9 | 146 | 14790.50 | 759417 | 2.41E-04 | 4 |
| 12F | 300 | 10 | 253 | 6725.08 | 1435544 | 2.09E-04 | 4.9 |
| 7F | 108 | 9 | 97 | 15037.27 | 591341 | 1.83E-04 | 5.5 |
| 15C | 151 | 10 | 146 | 4918.19 | 795484 | 1.90E-04 | 5.9 |
| 3 | 1421 | 10 | 1060 | 4855.85 | 6312025 | 2.25E-04 | 6.2 |
| 15B | 182 | 10 | 173 | 4681.43 | 969397 | 1.88E-04 | 7.7 |
| 9V | 15 | 7 | 15 | 4784.47 | 64999 | 2.31E-04 | 7.9 |
| 17F | 120 | 10 | 116 | 4732.12 | 554472 | 2.16E-04 | 8.3 |
| 11A | 383 | 10 | 355 | 4689.81 | 1913428 | 2.00E-04 | 8.9 |
| 6A | 13 | 6 | 13 | 5265.90 | 66380 | 1.96E-04 | 9.3 |
| 38 | 138 | 10 | 130 | 3635.66 | 793945 | 1.74E-04 | 12 |
| 23A | 487 | 10 | 437 | 4465.81 | 2450863 | 1.99E-04 | 12.2 |
| 8 | 416 | 10 | 339 | 4162.81 | 1886683 | 2.20E-04 | 12.2 |
| 10A | 189 | 10 | 181 | 4966.21 | 997469 | 1.89E-04 | 12.5 |
| 20 | 295 | 10 | 257 | 5671.61 | 1362075 | 2.17E-04 | 12.5 |
| 19A | 367 | 10 | 341 | 5227.81 | 1788357 | 2.05E-04 | 12.6 |
| 9N | 565 | 10 | 499 | 4571.55 | 2774894 | 2.04E-04 | 13 |
| 19F | 346 | 10 | 325 | 4898.12 | 1838106 | 1.88E-04 | 13.2 |
| 22F | 1035 | 10 | 854 | 4347.90 | 4998104 | 2.07E-04 | 13.2 |
| 18C | 7 | 4 | 7 | 3771.06 | 30961 | 2.26E-04 | 13.5 |
| 14 | 13 | 6 | 13 | 9481.60 | 69620 | 1.87E-04 | 14 |
| 16F | 385 | 10 | 359 | 4729.83 | 1873414 | 2.06E-04 | 15 |
| 21 | 40 | 10 | 40 | 4131.21 | 220528 | 1.81E-04 | 15.4 |
| 33F | 436 | 10 | 390 | 3406.75 | 2235386 | 1.95E-04 | 16.3 |
| 23F | 6 | 4 | 6 | 1224.26 | 25338 | 2.37E-04 | 16.7 |
| 15A | 462 | 10 | 423 | 4362.58 | 2258447 | 2.05E-04 | 17.3 |
| 6B | 19 | 8 | 19 | 5860.13 | 94637 | 2.01E-04 | 19.9 |
| 35F | 152 | 10 | 149 | 4434.52 | 809622 | 1.88E-04 | 31.9 |

**Table S2:**
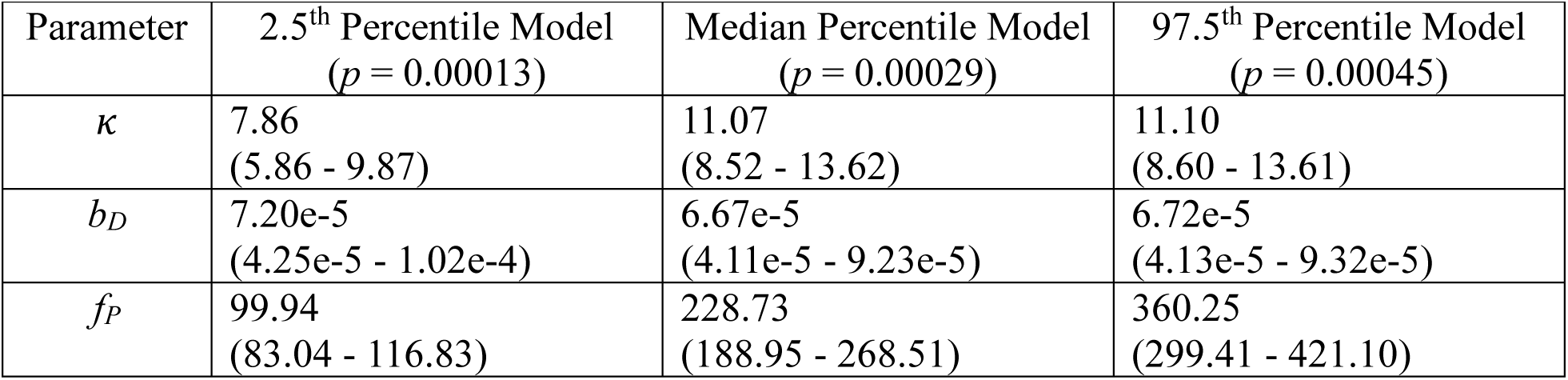
Piecewise model parameter estimates across varying fixed levels of invasiveness parameter *p*. The 95% confidence intervals are shown in parenthesis.

